# Additional Evidence Implicating GPD1L in the Pathogenesis of Brugada Syndrome in A Large Multi-generational Family

**DOI:** 10.1101/2022.09.17.22280058

**Authors:** Alexander Greiner, Haider Mehdi, Chloe Cevan, Rebecca Gutmann, Barry London

**Author notes:** Address for correspondence: Barry London, M.D., Ph.D., 285 Newton Road, 2283 Carver Biomedical Research Building, Iowa City, Iowa 52242, United States of America.

## Abstract

**Background:** Brugada Syndrome (BrS) is an inherited arrhythmia syndrome in which mutations in *SCN5A* account for 20% of cases. Mutations in other ion channels or channel-modifying genes may account for an additional 10% of cases, though recent analysis has suggested that *SCN5A* should be regarded as the sole monogenic cause of BrS.

**Objective:** We sought to re-assess the genetic underpinnings of BrS in a large mutligenerational family with a putative GPD1L-A280V mutation.

**Methods:** Fine linkage mapping was performed in the family using the Illumina Global Screening array. Whole exome sequencing of the proband was performed to identify rare variants and mutations, and Sanger sequencing was used to assay previously-reported risk single nucleotide polymorphsims (SNPs) for BrS.

**Results:** Linkage analysis decreased the size of the previously-reported microsatellite linkage region to ∼3 megabases. GPD1L-A280V was the only rare coding non-synonymous variation present at *<*1% allele frequency in the proband within the linkage region. Other variants were either synonymous, or in genes not known to play a role in BrS and that failed to co-segregate with BrS in the large family. Risk SNPs known to predispose to BrS were overrepresented in affected members of the family.

**Conclusion:** Together, our linkage and sequencing data suggest GPD1L-A280V remains the most likely cause of BrS in this large mutligenerational family. While care should be taken in interpreting variant pathogenicity given the genetic uncertainty of BrS, our data support inclusion of other putative BrS in clinical genetic panels.

## 1 Introduction

Brugada Syndrome (BrS) is an inherited arrhythmia syndrome characterized by ST-segment elevation in the right precordial leads (V_1_ through V_3_) and sudden cardiac death.^1^ The molecular mechanism underpinning BrS was initially described as autosomal-dominant loss-of-function mutations in the main cardiac sodium channel, Na_V_1.5, encoded on chromosome 3 by *SCN5A*.^2^ These loss-of-function mutations result in decreased inward depolarizing sodium current, which can result in premature repolarization of the epicardium in the right ventricle, slowed conduction, ventricular tachyarrhythmias and sudden cardiac death.^3^ However, only around 20% of patients with Brugada Syndrome have mutations in *SCN5A*. The advent of massively parallel sequencing has allowed many groups to investigate the genetic underpinnings of BrS over the past two decades. These genetic data, in combination with molecular studies, have expanded the number of putative disease-causing genes in BrS from one gene, *SCN5A*, to over 20 genes today. Genes reported to have mutations which cause BrS can be broadly classified as those which encode 1) sodium channels, 2) potassium channels, 3) channel interacting proteins, and 4) metabolic proteins.^4^ Recent reports have also suggested BrS may be polygenic in nature, with single nucleotide polymorphisms (SNPs) at the SCN5A-SCN10A and HEY2 loci predisposing individuals to BrS.^5^

Our lab first reported a large, multigenerational family with BrS.^6^ Initial linkage mapping using microsatellites identified an ∼15 centimorgan (CM) region on chromosome 3 with a LOD score *>*4 that did not contain *SCN5A* or *SCN10A*.^7^ Positional cloning and subsequent molecular analysis identified a variant in glycerol 3-phosphate dehydrogenase 1-like (GPD1L) as the putative cause of this family’s disease. Peak sodium current in HEK293 cells transfected with Na_V_1.5 decreased if cells were co-transfected with GPD1L-A280V compared to those co-transfected with GPD1L-WT. Additional studies suggested GPD1L-A280V decreases Na_V_1.5 membrane expression. Together, a decrease in peak sodium current and a decrease in membrane localization suggest that GPD1L-A280V is pathogenic for BrS.

The explosion of genetic data, especially variants of uncertain significance, from clinical genetic testing has led to a plethora of putative mutations but a dearth of scientific evidence supporting the pathogenicity of these variants. Concerns about variants of uncertain significance being classified as pathogenic without data to support the claim has been reviewed in many publications.^8,9^ This concern sparked a review of the pathogenicity of reported BrS genes using the Clinical Genome Resources (ClinGen) framework. This review determined that *SCN5A* was the only gene with sufficient scientific evidence to definitively be regarded as a causative gene for BrS.^10^ *GPD1L* was excluded due to a large linkage region in which other genes had not been sequenced and because the allele frequency of GPD1L-A280V was relatively high (gnomAD V2.1.1 allele frequency = 1.29E-4).^11^

In the present study, we provide high-depth whole exome sequencing data in the proband of our originally-reported large multigenerational family, SNP-based linkage analysis of affected individuals, and sequencing data of previously defined BrS risk SNPs to support our initial reports that GPD1L-A280V is pathogenic for BrS.

## 2 Methods

### 2.1 Patient enrollment and phenotype validation

Patients were enrolled under protocols approved through the University of Pittsburgh and University of Iowa Institutional Review Boards (IRBs). All individuals in the study provided informed consent for participation in the study. Type 1, Type 2, or Type 3 Brugada Syndrome, or clinically unaffected status, was determined through clinical history, electrocardiogram analysis, and clinical provocative testing using procainamide as previously described.^6^

### 2.2 Genomic DNA isolation and RNAse A treatment

Genomic DNA was isolated from peripheral blood using commercially-available kits or automated machine as previously described.^6^ Isolated DNA was treated with RNAse A and re-isolated by ethanol precipitation prior to sequencing. DNA integrity was assessed by agarose gel electrophoresis and quantified using Qubit High Sensitivty DNA assays (ThermoFisher Scientific).

### 2.3 SNP calling and Linkage Analysis

Ten affected and five unaffected individuals from the family had genomic DNA prepared as described above. After passing quality control and subsequent library preparation, SNPs were called using the Infinium Global Screening Array (Illumina) at CD Genomics (Shirley, NY, USA) which sequences approximately 654,027 SNPs. Call rates were greater than 98% for all individuals. (Supplemental Table 1) Chromosomes were phased using ShapeIt2 with DuoHMM enabled and the reference HapMap Phase II data.^12,13^ The resultant phased haplotypes were investigated manually for recombination using our previously-reported microstaellite data as the starting point. Logarithm of odds scores were calculated using Superlink-Online SNP.^14^

### 2.4 High-depth whole exome sequencing and analysis

Whole exome sequencing was performed at the Iowa Institute of Human Genetics (IIHG) on genomic DNA prepared as described above. Library preparation was performed using standard protocols for the Agilent SureSelect V6 + UTR library preparation kit (Agilent Technologies). The resulting 150 base pair paired-end library was sequenced on an Illumina HiSeq 4000. Resultant .fastq files underwent quality control using FastQC.^15^ Alignment, variant calling, and variant annotation were performed using BWA-MEM (alignment), GATK4 (variant calling), and SnpEff (variant annotation) using an implementation of BCBIO and the GRCh37 reference genome.^16–21^. A second variant calling tool, Freebayes, was used in a near-identical pipeline to ensure consensus variant calls were achieved^22^. Greater than 80% of targeted bases had greater than 50x coverage. (Supplemental Figure 1) A brief summary of sequencing results is shown in Supplemental Table 2.

Two approaches were used to identify candidate variants 1) a linkage-region focused analyses and 2) a linkage-naive approach. The linkage-region focused analysis was limited to the newly-defined linkage region on chromosome 3, while the linkage-naive approach entertained variants on any autosome. For both approaches, the resulting annotated variant call file was a used to generate a database compatible with Gemini.^23^ Variants present at less than 1% minor allele frequency, a lenient threshold for autosomal dominant disease, were included. Variants were then segregated according to impact (nonsense, splicing, missense, synonymous) with synonymous variants removed from consideration. Variant location information, either exonic, UTR, intronic, or intergenic, was subsequently used in prioritization of variants.

### 2.5 Sanger Sequencing of Risk SNPs

Sanger sequencing of BrS risk SNPs was performed at the Iowa Institute of Human Genetics using custom primers designed in-lab and synthesized by Integrated DNA Technologies. (Supplemental Table 3) The resulting chromatograms were analyzed using NCBI Blast, Finch, and SnapGene from which wild-type, heterozygous, or homozygous SNP calls were made and manually validated.

### 2.6 Protein and variant modeling

A single unpublished protein model of GPD1L is present on the RCSB Protein Databank.^23,24^. AlphaFold computational models for GPD1L are also publicly available from EMBL. The AlphaFold GPD1L model AF-Q8N335-F1-model v1 was downloaded and imported into PyMol.^25^ The mutagenesis function of PyMol was used to visualize the impact of the three predicted GPD1L-A280V rotamers. Strain scores were calculated for all rotamers within PyMol. (Supplemental Figure 2) Variant data for *GPD1L* was downloaded from ClinVar.^26^

## 3 Results

### 3.1 Linkage analysis reaffirms a region on chromosome 3 co-segregates with Brugada Syndrome

To understand the linked genomic regions within this family we performed linkage analysis using the Illumina Global Screening Array with ten affected and five unaffected family members. The previously-identified 15 CM linkage region in this family which we reported was used as a starting point for our investigation.^6^ Analysis of phased haplotypes identified a narrowed linkage region defined by the SNPs rs13059657 and rs7651953 (GRCh37, chr3:29,899,567-32,970,737). (Figure 1) Individuals III-2 and III-8 defined the 5’ and 3’ breakpoints, respectively. Calculation of LOD scores using individuals having a Type 1 EKG pattern or positive procainamide test, in addition to I-I and I-II from Figure 1, using Superlink-Online SNP results in a LOD score of 2.02 for this region.^14^ The addition of individuals with Type II EKG patterns results in a LOD score of 3.18. The region of interest contains 14 genes, including *GPD1L*, but not *SCN5A* or *SCN10A* which are approximately 6 Mb downstream. (Figure 1)

**Figure 1:**
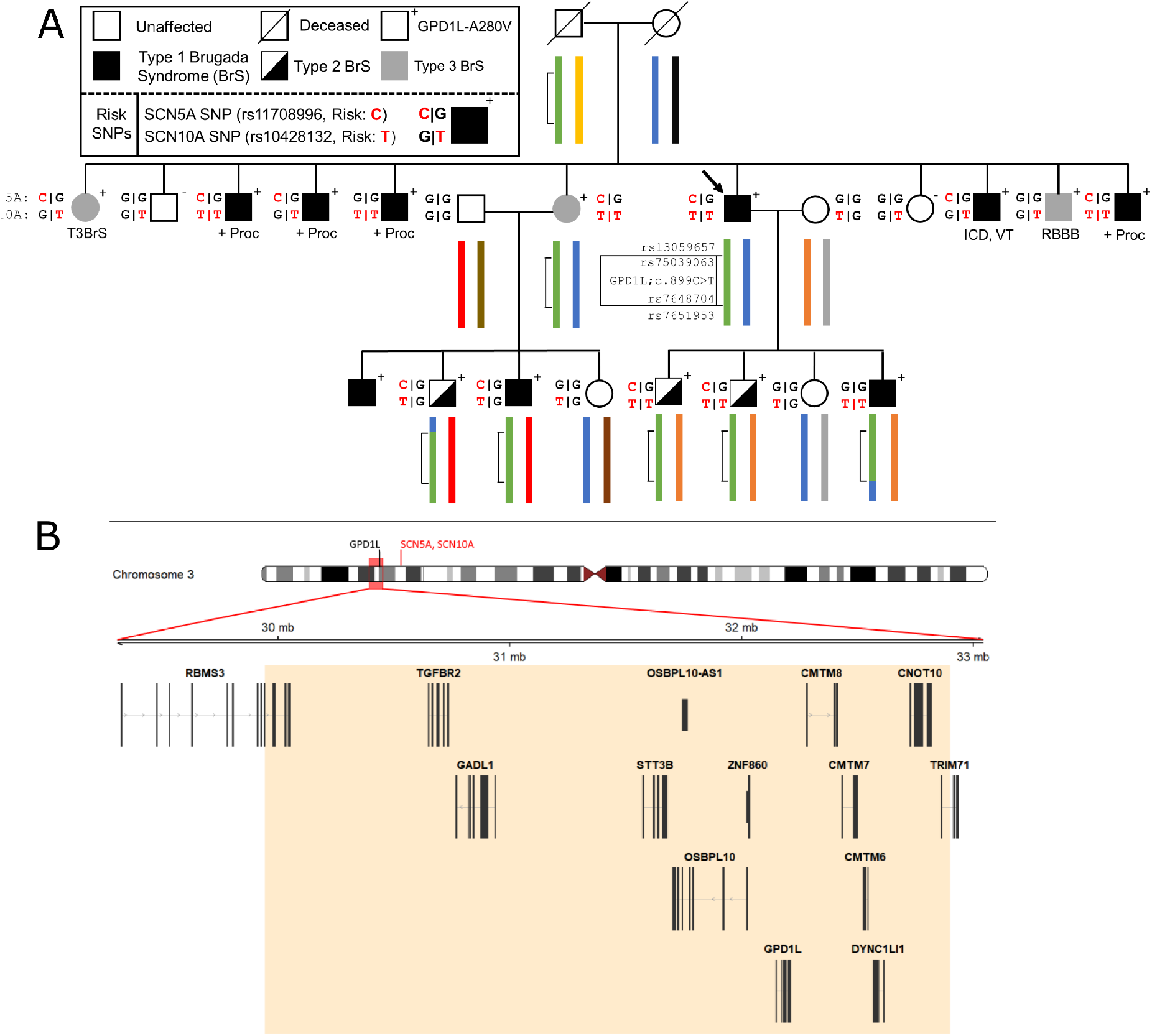
Representative pedigree of the family, linkage analysis, and assessment of Brugada Syndrome (BrS) risk SNPs. A) Linkage analysis demonstrates a small region on chromosome 3 (inset map and green brack-eted region) which has undergone recombination in the third generation of the pedigree. Previously reported risk SNPs (SCN5A and SCN10A, red) for the BrS phenotype are associated with the GPD1L-A280V. B) The 3 megabase region on chromosome 3 identified by linkage analysis contains 14 genes, including *GPD1L. SCN5A* and *SCN10A* lie upstream of GPD1L (ideogram, red text) and outside of the refined linkage region (ideogram, red box). ICD: implantable cardioverter defibrillator. +Proc: positive procainamide challenge. RBBB: right bundle branch block. VT: ventricular tachycardia.

### 3.2 Whole-exome sequencing analysis affirms GPD1L-A280V as the only rare exonic variant in the linked region

High-depth (100x) whole exome sequencing of the proband was performed using the Agilent SureSelect V6+UTR capture kit. Limiting the analysis to the newly defined linkage region identified GPD1L-A280V as the only rare, exonic coding variation. No other exonic or splice variants occurred at an allele frequency of less than 1%. The proband carried missense variants in arrhythmia and cardiomyopathy genes not usually associated with BrS: one missense variant in ANK2 (p.R2416G;c.7246C*>*G, unreported), one in FHL2 (p.V187M;c.559G*>*A, f= 6.78E-5) and two in TTN (p.A31503T;c94507G*>*A, f= 5.04E-5 and p.F14410C;c.43229T*>*G, unreported) (Table 1). Linkage analysis and Sanger sequencing showed that none of these variants cosegregate with BrS in this family, and no other variants are annotated as pathogenic for cardiovascular diseases in the ClinVar database.

**Table 1.** Variants in reported Brugada Syndrome or Inherited Cardiac Condition (ICC) List Genes

|  |  |  |  |  |
| --- | --- | --- | --- | --- |
| Gene List | Position | Consequence | Gene | Frequency |
| Linkage region | chr3:32200588C>T | p.Ala280Val | GPD1L | 1.29E-04 |
| BrS | chr12:2788732C>T | p.Gly1738Gly | CACNA1C | 1.21E-05 |
| ICC | chr2:105979871C>T | p.Val187Met | FHL2 | 6.78E-05 |
|  | chr2:179411745C>T | p.Ala31503Thr | TTN | 5.04E-05 |
|  | chr2:179497504A>C | p.Phe14410Cys | TTN | Unreported |
|  | chr4:114277020C>G | p.Arg2416Gly | ANK2 | Unreported |
|  | chr16:15844004G>A | p.His690His | MYH11 | 5.25E-04 |

### 3.3 The SCN5A and SCN10A BrS risk SNPs are linked to GPD1L-A280V in this family

Three published risk SNPs for BrS [rs10428132 (*SCN10A*; total minor allele frequency, dbSNP (MAF) = 0.40), rs11708996 (*SCN5A*; MAF=0.14), and rs9388451 (*HEY2* ; MAF=0.42)] identified by Bezzina et al. were assessed in family members with sufficient DNA, showing 8/15 affected subjects (those demonstrating a Type I, II, or III BrS pattern) in generations II and III are homozygous for the SCN10A risk allele and 12/15 subjects are heterozygous for the SCN5A risk allele (Figure 1). SCN5A and SCN10A are near GPD1L on chromosome 3; in most affected individuals the GPD1L-A280V mutation co-segregated with one SCN5A and one SCN10A risk allele. The HEY2 risk SNP on chromosome 6 segregated in Mendelian fashion. Overall, individuals affected with BrS (n=15) carried 3.3 ±0.2 risk SNPs, while unaffected individuals (n=12) carried 2.1 ±0.4 risk SNPs (p=0.012). (Figure 2) The SCN5A and SCN10A risk SNPs carried along with GPD1L-A280V in this family may potentiate the pathogenicity of GPD1L-A280V.

**Figure 2:**
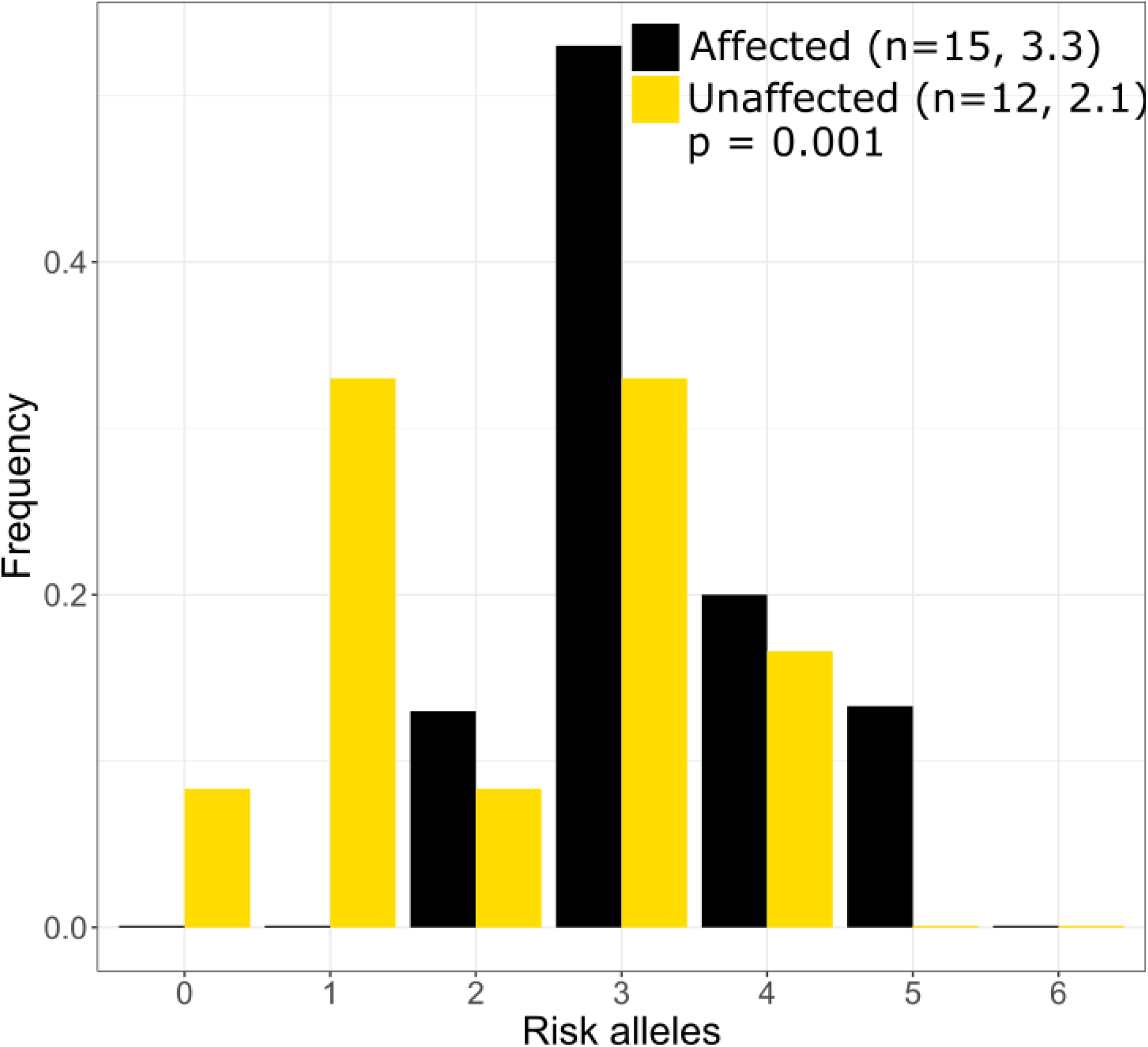
Distribution of Brugada Syndrome Risk Polymoprhisms in the large mutli-generational family. Individuals with Brguada Syndrome in our family carry more risk polymorphisms on average (3.3) than unaffected individuals (2.1) in our large mutli-generational family. The distribution of risk poly-moprhisms is similar to that which was reported by Bezzina et al.^5^

### 3.4 GPD1L-A280V may introduce protein instability in computationally-predicted models

*In silico* mutagenesis of the A280 residue to V280 in the computational model of GPD1L demonstrates increased strain within the alpha helix in which residue 280 resides. (Figure 3 and Supplemental Figure 2) Thus, *in silico* mutagenesis results in increased computationally-prediced strain stores indicating GPD1L-A280V may introduce steric hinderance in the GPD1L tertiary structure.

**Figure 3:**
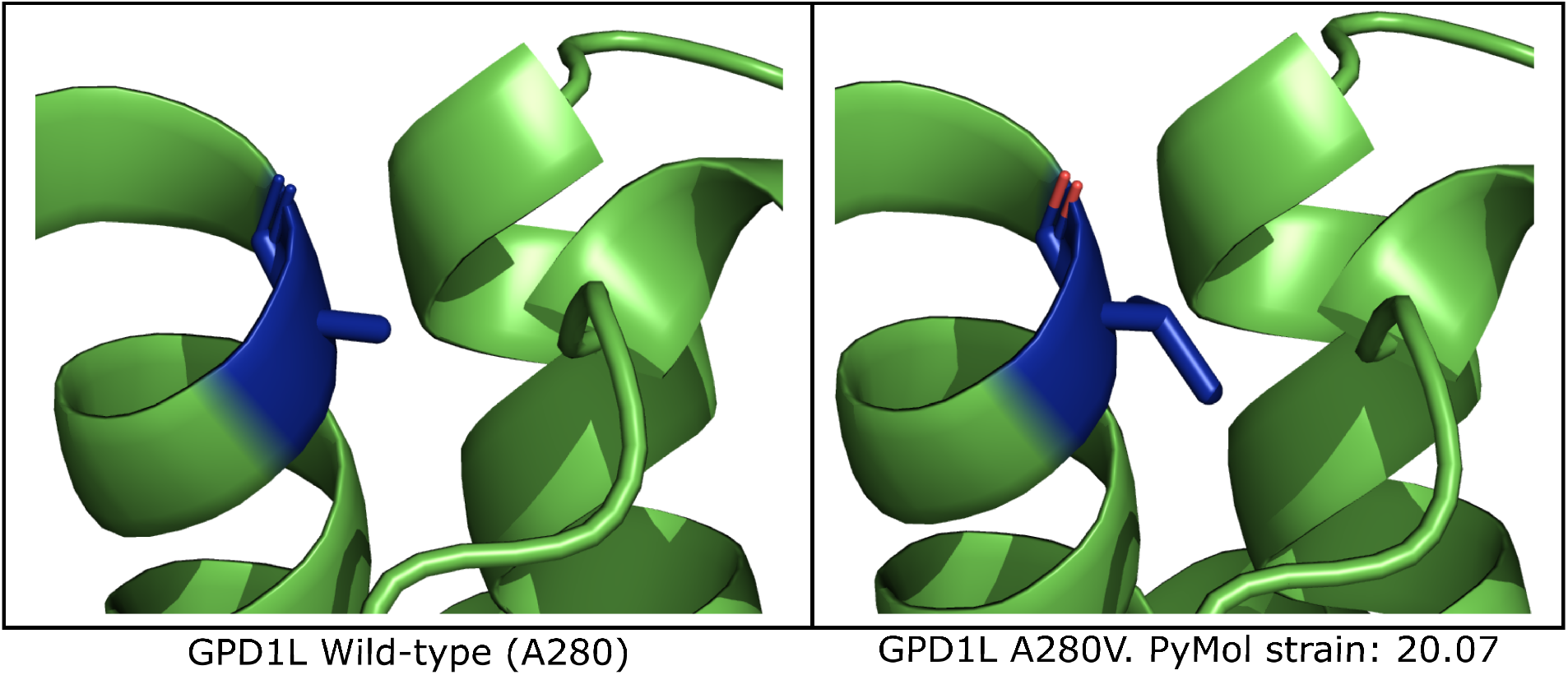
Computational modeling of GPD1L suggests A280V introduces steric clashing. Left: The wild-type GPD1L structure as calculated by AlphaFold (AF-Q8N335-F1-model v1) with residue A280 highlighted in blue. A280 resides in an alpha helix. Right: One of three putative GPD1L-A280V rotamers as calculated by PyMol. All three predicted rotamers introduce strain in the alpha helix.

## 4 Discussion

### 4.1 The genetic architecture of BrS in one large, multigenerational family

Our analysis of this large, multigenerational family narrows the linkage region to approximately 3.1 megabases which includes *GPD1L* but excludes *SCN5A* and *SCN10A*. Whole exome sequencing confirmed GPD1L-A280V as the only rare exonic variant at an allele frequency *<*1% within this region. The affected-only LOD score of 3.18 for individuals displaying Type I and Type II BrS patterns (Type 1 BrS alone = 2.02), suggests this region is linked to BrS in this family and is consistent with the previous reportd LOD score of *>*3 which included unaffected individuals. While not proving pathogenicity of the GPD1L-A280V mutation, these data, along with previously published experiment data should be considered when assessing the role of GPD1L in the pathogenesis of BrS.

While the allele frequency of GPD1L-A280V is high relative to published allele frequency cutoffs, GPD1L-A280V is not the lone reported *GPD1L* putative mutation.^27^ Four other putative mutations, p.I124V, p.E174K, p.R231C, and p.Q345H, have been reported in ClinVar. (Figure 4)^28^ Nevertheless, we believe that mutations in GPD1L, in isolation, are likely unable to cause sufficient decreases in sodium current to precipitate BrS. It is likely that GPD1L mutations and risk polymorphisms act in concert to precipitate BrS, as is the case of this large multigenerational family. We believe that SCN5A is the only gene in which mutations are sufficient to cause enough inward sodium current decrease to cause BrS, while GPD1L —and possibly other reported BrS genes —precipitate BrS in a polygenic manner or alongside risk SNPs.

**Figure 4:**
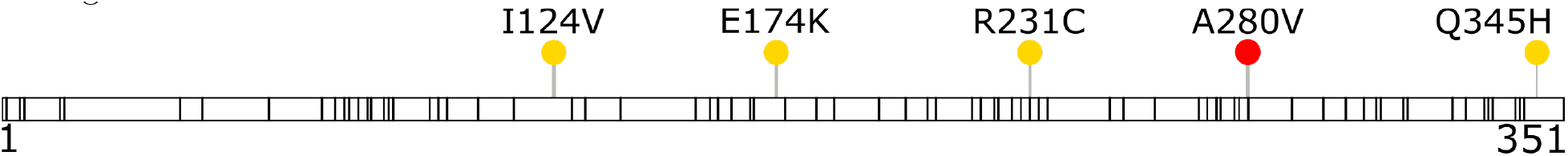
Reported GPD1L variants in ClinVar show GPD1L-A280V is not the sole reported putative mutation. Missense variants with conflicting interpretation (yellow circles, with GPD1L-A280V highlighted as red) and variants of uncertain significance (black bars, 83 total) were downloaded from ClinVar (accessed November 12, 2021) and plotted along the full length of GPD1L. All five variants with conflicting interpre-tation, including GPD1L-A280V, were submitted to ClinVar by their respective submitters with Brugada Syndrome as the clinical condition.

### 4.2 The genetic architecture of BrS: past, present, and future

Mutations in *SCN5A* are certainly the predominant genetically-identifiable cause of BrS.^1^ Future studies of *GPD1L* and other putative BrS mutations in heterologous systems and in mouse models will continue provide information on the role these genes play in cardiac physiology. Surprisingly, few reports exist of the use of whole-exome or whole-genome sequencing to identify the genetic underpinnings of BrS, and the majority of these studies identify genetic variation in previously reported disease genes. Future large genomic datasets (e.g. UK Biobank, gnomAD) have and will continue to provide further information on allele frequency of genes reported to cause BrS. In all, these data and future studies will further our understanding of the still uncertain genetic architecture of BrS.

### 4.3 Study limitations

Our study uses linkage and exome sequencing data from a single family or single proband. Linkage data provide an incomplete picture of the genetic architecture for a disease, though we believe the addition of exome sequencing partly addresses this limitation. Notably, exome sequencing libraries are not targeted to capture intronic sequences. Deep intronic variants within *GPD1L* may contribute to disease in this family.

### 4.4 Clinical implications

Recent recommendations to clinical genetic testing panels exclude all genes except *SCN5A* in their testing panels. While well-intentioned to avoid undue harm from provider misinterpretation, this may reduce identification of rare variation contributing to BrS in sodium channel modifying genes, among others. While concerns for generating undue harm from misinterpretation of a variant of uncertain significance in a disputed gene included on a clinical genetic testing panel are valid, we believe these concerns highlight the necessity of thorough understanding of genetic testing results and the importance both of cardiologists trained in genetics and the use of genetic counselors.^29,30^

Our study demonstrates that mutation in GPD1L likely contributes to BrS in this multigenerational family. Given this, inclusion of *GDP1L* on genetic testing panels should be reconsidered.

## 5 Conclusion

We identified GPD1L-A280V as the only rare non-synonymous coding variation in the refined linkage region in a large, multigenerational family. Associated BrS risk SNPs may be permissive for the BrS phenotype in affected family members. In addition, the A280V mutation may alter the GPD1L tertiary structure. While mutation in *SCN5A* is the most common cause genetically-identified cause of BrS, our study suggests that GPD1L should still be considered when evaluating the genetic underpinnings of BrS.

## Data Availability

All data produced in the present study are available upon reasonable request to the authors

## Abbreviations

BrS: Brugada Syndrome
SNP: Single nucleotide polymorphism
CM: centimorgan
LOD: logarithm of the odds
Mb: megabase
ICD: implantable cardioverter defibrillator
RBBB: right bundle branch block
VT: ventricular tachycardia

