## SupplementalData for "Additional Evidence Implicating GPD1L in the Pathogenesis of Brugada Syndrome in A Large Multi-generational Family"

Supplemental Table 1: Illumina Global Sequencing Array Call Rate

| **Sample** | **Call Rate** |
|---|---|
| 1 | 0.9955 |
| 2 | 0.9937 |
| 3 | 0.9955 |
| 4 | 0.9950 |
| 5 | 0.9948 |
| 6 | 0.9953 |
| 7 | 0.9955 |
| 8 | 0.9954 |
| 9 | 0.9955 |
| 10 | 0.9919 |
| 11 | 0.9923 |
| 12 | 0.9865 |
| 13 | 0.9938 |
| 14 | 0.9954 |
| 15 | 0.9955 |

Supplemental Figure 1: **Whole exome sequencing coverage**. Good coverage was achieved for the Agilent SureSelect V6+UTR kit, with >80% of bases covered above 50x.

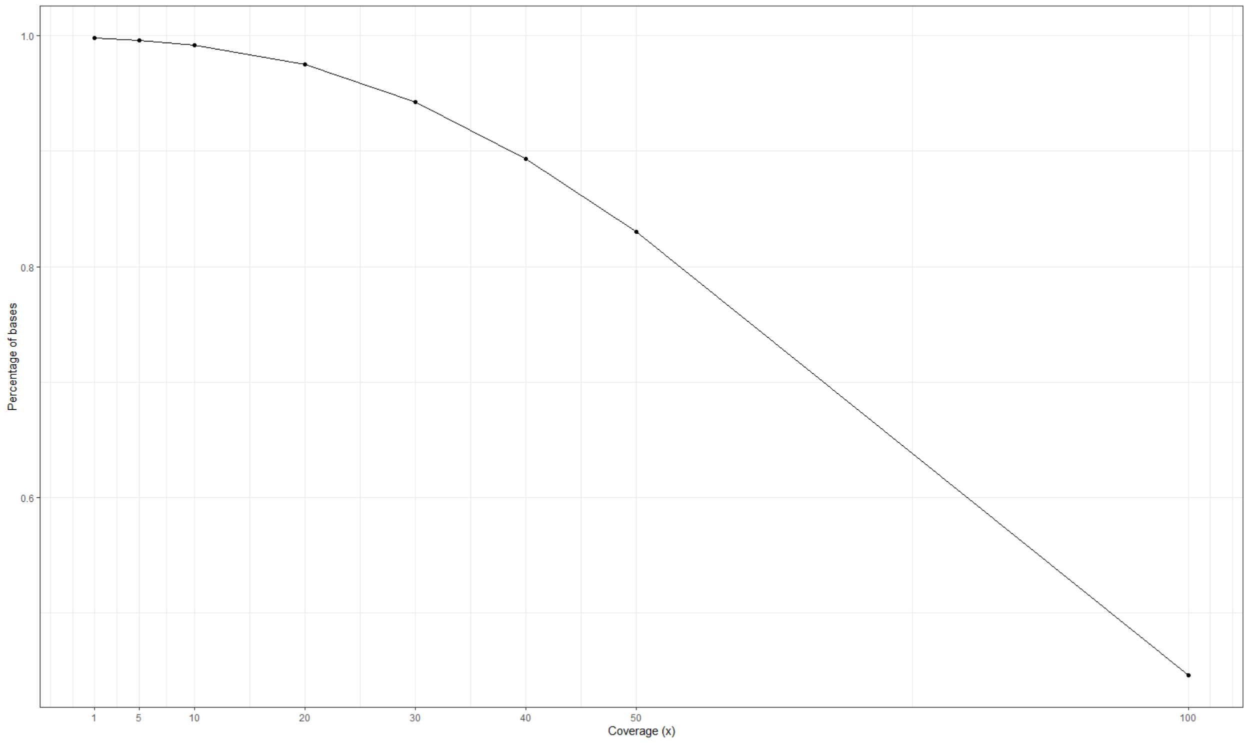

Supplemental Table 2: Whole exome sequencing metrics

| | |
|---|---|
| **Number of variants** | 166834 |

**Variants by type**

| Type | Count | Percent |
|---|---|---|
| DEL | 14624 | 8.77 |
| INS | 12256 | 7.35 |
| SNP | 139954 | 83.89 |

**Variants by effect**

| Effect | Count | Percent |
|---|---|---|
| MISSENSE | 34535 | 47.26 |
| NONSENSE | 308 | 0.42 |
| SILENT | 38238 | 52.32 |

Supplemental Table 3: Primers used for Sanger Sequencing of Brugada Syndrome Risk SNPs

| Primer | Gene | Sequence |
|---|---|---|
| rs11708996 F1 | SCN5A | TGTTGAGTTGTAGGGTACACAGT |
| rs11708996 R1 | SCN5A | GGGATTGGGAAGGCCTTCAA |
| rs11708996 F2 | SCN5A | ATGTTGATTCCAGTTTCCCCT |
| rs11708996 R2 | SCN5A | TGAACTCACTACCACAAACTGGA |
| rs10428132 F1 | SCN10A | CAGAGCAAATGGAGCAAGGC |
| rs10428132 R1 | SCN10A | CCAGTTCACCAGTCTCCGTC |
| rs9388451 F1 | HEY2 | CGAGGTGCCAGGGGTTTTTA |
| rs9388451 R1 | HEY2 | GCGAGGAGAATACCAGAGGC |
| rs9388451 F2 | HEY2 | CCACAGTGACATGATCCAGGT |
| rs9388451 R2 | HEY2 | GGAACATGTGCAAGGCCTTG |

Supplemental Figure 2: **GPD1L-A280V PyMol Rotamers**. The AlphaFold predicted model for GPD1L was mutated in PyMol to GPD1L-A280V. Red octagons indicate steric hinderance within GPD1L. A) PyMol predicted rotamer 1 of 3 with a calculated strain of of 14.29. B) PyMol predicated rotamer 2 of 3 with a calculated strain of 14.63. C) PyMol predicted rotamer 3 of 3 20.07

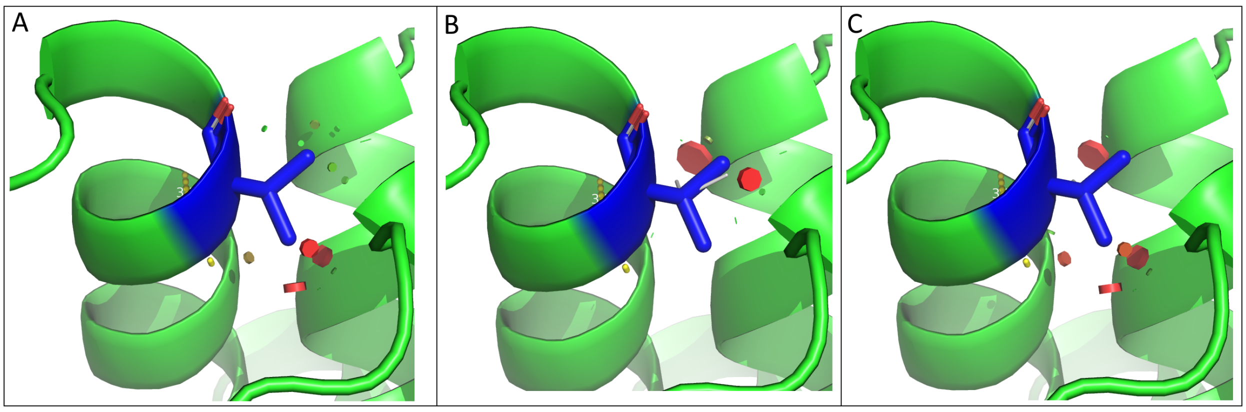
